# Use of real-world evidence data to evaluate the comparative effectiveness of second-line type 2 diabetes medications on chronic kidney disease

**DOI:** 10.1101/2021.06.15.21258963

**Authors:** Yu Deng, Farhad Ghamsari, Alice Lu, Jingzhi Yu, Lihui Zhao, Abel N Kho

**Author notes:** **Corresponding author**: Yu Deng, BS, Center for health information Partnerships (CHIP), Northwestern University Feinberg School of Medicine, 625 N. Michigan Ave, Suite 1500, Chicago, IL 60611. **Funding source**: not applicable.

## Abstract

Chronic kidney disease (CKD) is a common complication of type 2 diabetes mellitus (T2DM). Approximately one-third of patients with T2DM also have CKD. In clinical trial studies, several anti-diabetic medications (ADM) show evidence of preventing the progression of CKD. Biguanides (e.g., metformin) are widely accepted as the first line medication. However, the comparativeness effectiveness of second line ADMs on CKD outcomes in T2DM is unclear. In addition, results from clinical trials may not generalize into routine clinical practice. In this study, we aimed to investigate the association of second line ADMs with incident CKD and CKD hospitalization in T2DM patients using real-world data from electronic health records. Our study found that treatment with sodium-glucose cotransporter 2 (SGLT-2) inhibitors was significantly associated with a lower risk of CKD incidence in both primary analysis (hazard ratio, 0.43; 95% CI, [0.22;0.87]; p-value,0.02) (SU) as a second-line ADM. Treatment with a dipeptidyl peptidase 4 (DPP-4) inhibitor was significantly associated with lower CKD incidence (hazard ratio, 0.7; 95% CI, [0.53;0.96]; p-value, 0.03) and lower CKD hospitalization events (hazard ratio, 0.6; 95% CI, [0.37; 0.96]; p-value, 0.04) in the primary analysis. However, both associations were not significant in the sensitivity analysis. We did not observe significant association between use of GLP-1, TZD, insulin and CKD incidence or hospitalization compared to use of SU as the second-line ADM.

## Introduction

Type 2 diabetes is a major risk factor for chronic kidney disease (CKD) and is the leading cause of end stage renal disease (ESRD) with approximately 20% - 40% of patients developing diabetic nephropathy ^1^. Sulfonylureas (SU), dipeptidyl peptidase 4 (DPP-4) inhibitor, glucagon-like peptide 1 receptor agonists (GLP-1RA), sodium-glucose cotransporter 2 (SGLT-2), Thiazolidinediones (TZD), and Insulin are commonly used as second-line medication in addition to metformin. In the past decade, several randomized clinical trials have shown evidence of the newer anti-diabetic medications (ADMs) such as SGLT-2 and GLP-1RA in reducing risk for renal disease outcome compared to placebo^2–6^. However, there are few clinical trials (ongoing or finished) that directly compare the effectiveness of the newer AMDs to the older ones such as SU and DPP-4 inhibitor. Furthermore, patients in clinical trial studies are more compliant with therapy for a number of reasons (e.g. support from study staff), therefore, it is unclear how well results from clinical trials may apply to the general population in real clinical practice^7^. Electronic health records (EHRs), a major source of real-world evidence data, can facilitate the understanding of treatment effectiveness in clinical practice using patient level data from the routine operation of the healthcare system and complement evidence on the efficacy of medications from randomized controlled trials (RCTs) ^8^. Recently, a study using real-world data investigated the comparative effectiveness of SGLT-2 inhibitor, GLP-1RA, DPP-4 inhibitor, and SU in type 2 diabetics for preventing renal disease ^9^. This study was conducted within the Department of Veterans Affairs, which serves an older (mean age = 65.46), white (71.95%), male (94.49%) population. Therefore, additional research is required to compare the effectiveness on renal disease across the multitude of currently available drugs and drug classes. In this study, we compared the effect of commonly used second line anti-diabetic medications including SU, DPP-4 inhibitor, GLP-1RA, SGLT-2 inhibitor, TZD, and Insulin on renal outcomes using EHRs from a large integrated health delivery system.

## Methods

### Study population

The Northwestern Medicine Electronic Data Warehouse (NMEDW) is the primary data repository for all the medical records of patients who receive care within the Northwestern Medicine system ^10^. Established in 2007, the NMEDW contains records for over 3.8 million patients, with most EHR data going back to at least 2002, and with some billing claims data going back to 1998 or even earlier ^10^. We included patients who met the following criteria: 1) at least one prescription to an ADM 2) at least one diagnosis code for type 2 diabetes (see supplemental Table 1 for type 2 diabetes diagnosis codes)^11^ 3) no excluded diagnoses: pregnancy, type 1 diabetes mellitus 4) at least one year of records in the database before their first ADM exposure and 5) at least three years of continuous ADM prescription records after first ADM exposure. Given that the first SGLT-2 inhibitor was approved by FDA for use in the United States in March 2013, we only included patients who had their first ADM exposure after 2013-03-01. Figure 1 shows the flow chart of the patient selection process.

**Table 1.**
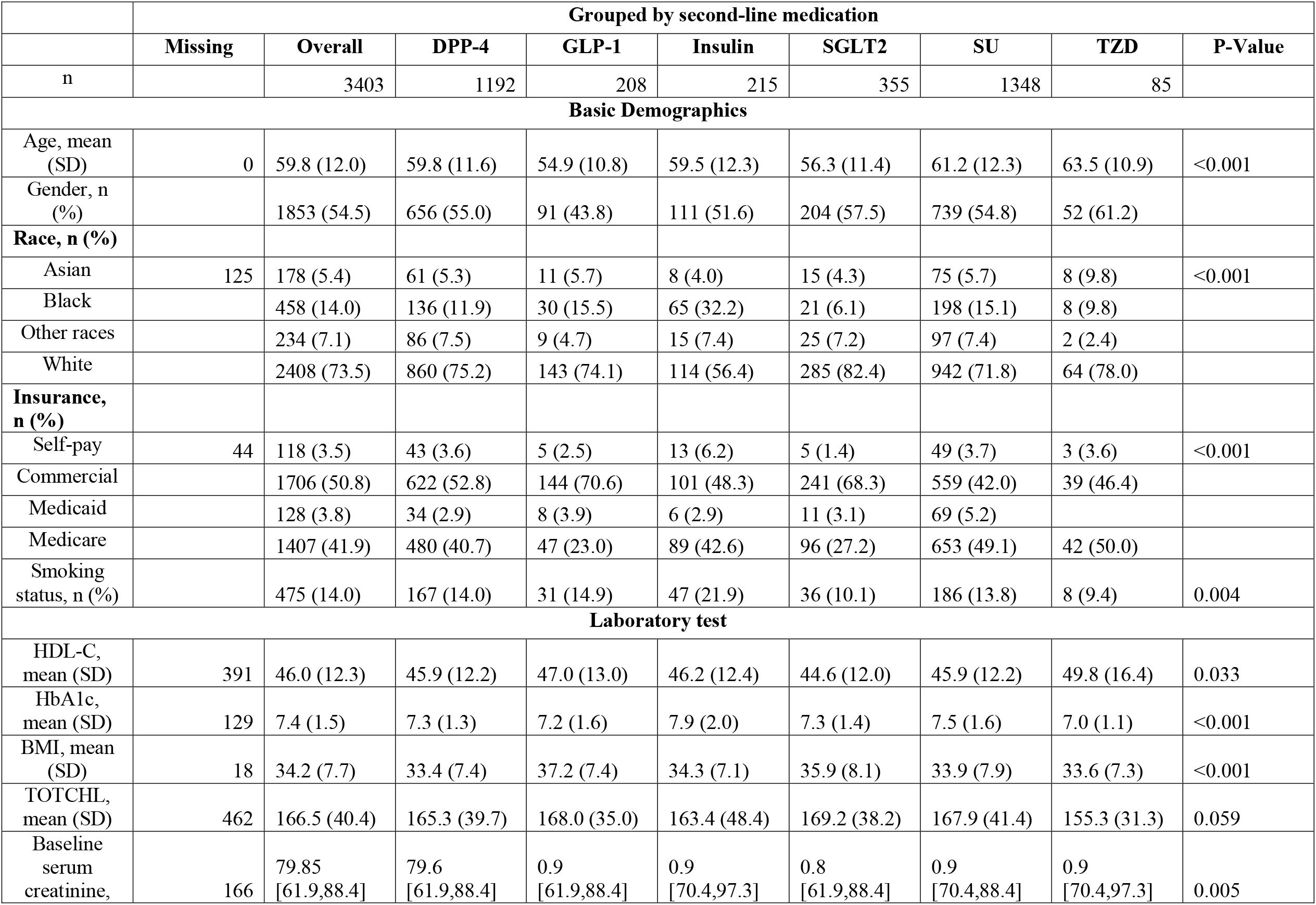
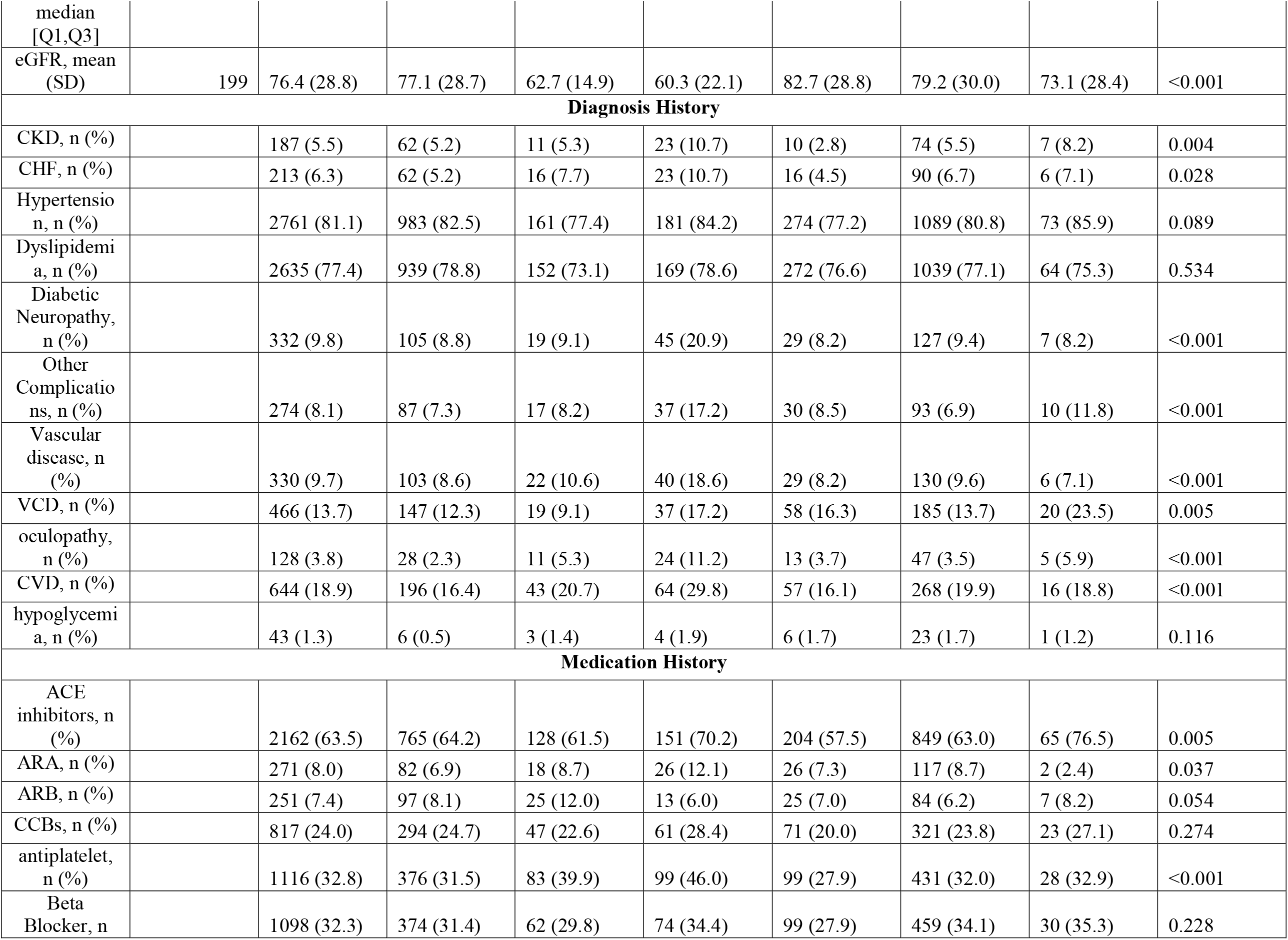
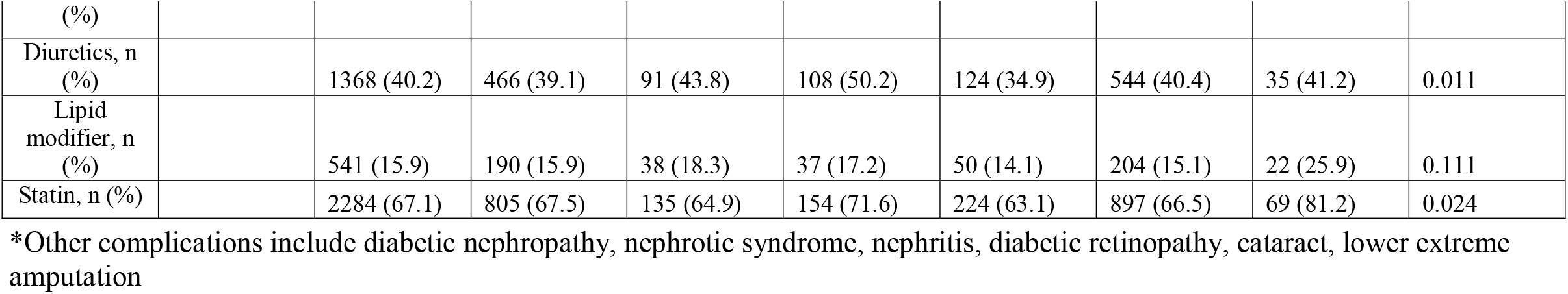
Baseline characteristics of study cohort. This table shows the baseline patient characteristics for CKD hospitalization outcome before missing data imputation. For CKD incidence outcome, patients with prior ESRD and prior CKD were excluded from the study cohort. For composite renal outcome, patients with prior ESRD were excluded from the study cohort. Abbreviations: HDL, high density cholesterol, HbA1c, hemoglobin A1c; BMI, body mass index; TOTCHL, total cholesterol; VCD: vascular complications of diabetes; CHF, congestive heart failure; CVD, cardiovascular disease; ACE inhibitor, angiotensin-converting enzyme inhibitors; ARA, aldosterone receptor antagonists; ARB, angiotensin II receptor blocker. DPP-4, dipeptidyl peptidase 4 inhibitors; GLP-1RA, glucagon-like peptide receptor agonists; SGLT-2, sodium-glucose cotransporter 2 inhibitor; TZD, Thiazolidinediones.

**Figure 1.**
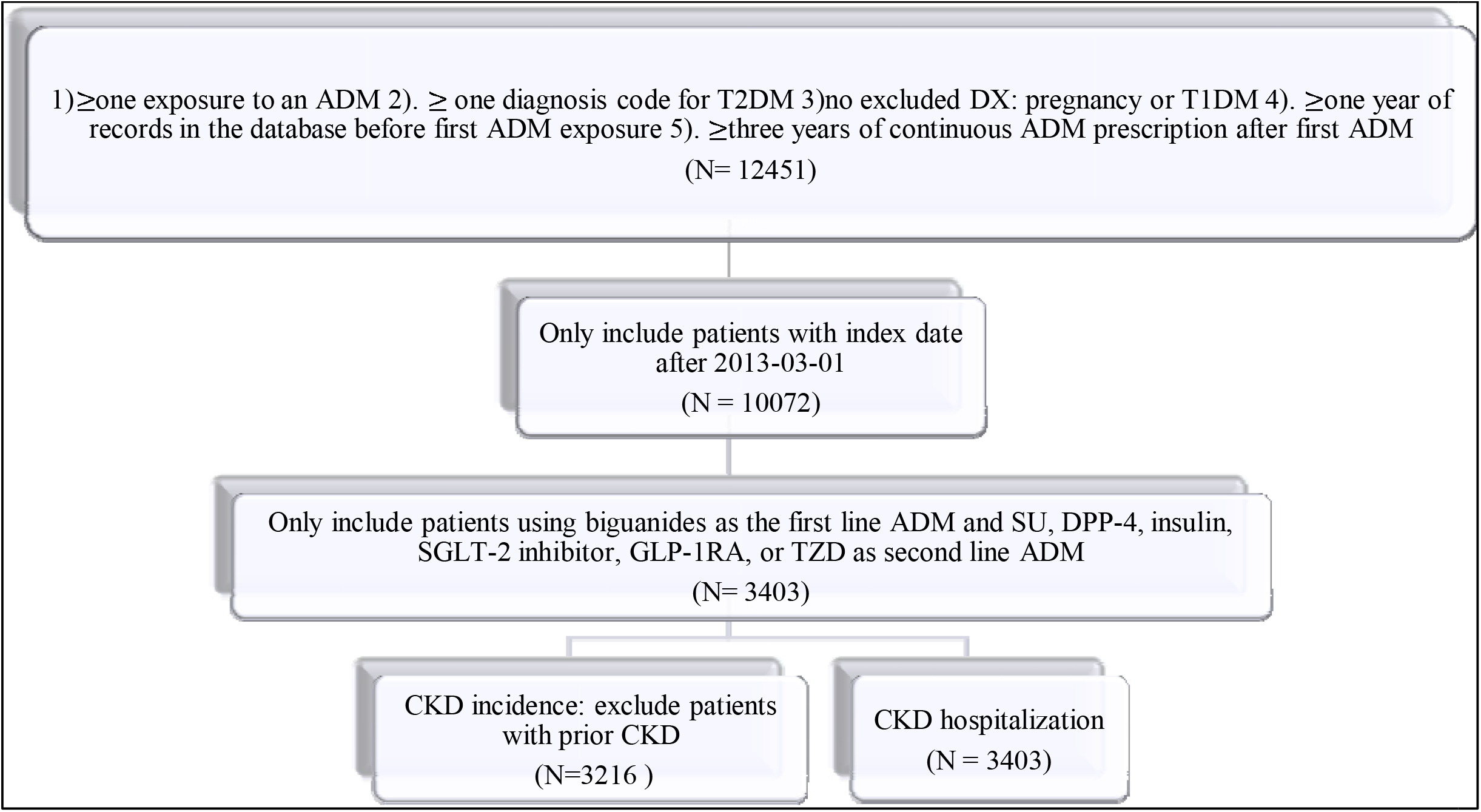
Cohort selection flowchart. Abbreviations: T2DM, type 2 diabetes mellitus; ADM, antidiabetic medication; DX, diagnosis; T1D, type 1 diabetes; SU, sulfonylureas; DPP-4 inhibitor, dipeptidyl peptidase 4 inhibitors; GLP-1RA, glucagon-like peptide receptor agonists; SGLT-2 inhibitor, sodium-glucose cotransporter 2 inhibitor; TZD, Thiazolidinediones.

We defined an ADM sequence as the chronological series of ADM prescription orders a patient received from the date of their first ADM prescription to one year later (e.g., “Metformin, Glipizide, Glipizide, Glucophage, Sitagliptin, Glipizide”). We then mapped each generic name and brand name to its respective drug class in Anatomical Therapeutic Chemical Classification (e.g., “Biguanides, SU, SU, Biguanides, DPP-4 inhibitor, SU”) ^12^. Repeated listings of the same medication in each sequence were understood to be refilled prescription. Therefore, in this example, only the first occurrence of a medication is kept in the final sequence (e.g., “Biguanides, SU, DPP-4 inhibitor”). During the 1-year period, only patients who were treated with biguanides as their first line of medication and had a second medication of either DPP-4 inhibitor, SGLT-2 inhibitor, GLP-1RA, SU, TZD, or Insulin were included in the study.

### Exposure

The exposure in this study was the medication sequence during a patient one-year exposure period as described above. The sequences we were interested in include ‘Biguanides, SU’, ‘Biguanides, DPP-4 inhibitor’, ‘Biguanides, Insulin’, ‘Biguanides, GLP-1RA’, ‘Biguanides, SGLT-2 inhibitor’ and ‘Biguanides, TZD’. A patient might switch to a third medication during the one-year exposure time, we considered them in the same group as the patients who did not. For example, ‘Biguanides, SU, DPP-4’ was considered in the same second-line ADM group as ‘Biguanides, SU’. Patients who were only on Biguanides during the 1-year drug exposure period were excluded from the study. Index date was defined as one year after first the exposure to ADM.

### Renal outcomes

Our primary outcome is CKD incidence identified by the first appearance of an associated International Classification of Diseases, 9^th^/10^th^ revision (ICD9/10) diagnosis code (see supplemental Table 1)^7^. Patients who have CKD, ESRD or macroalbuminuria before index date are excluded in the analysis for this outcome. In addition to the incident CKD event, we evaluated ADMs’ associations with CKD hospitalization. CKD hospitalization were identified based on ICD9/10 codes. For any of the outcomes, patients were followed up from their index date until meeting criteria of one of the above outcomes, last hospital visit date, end of 5-year follow up period or end of study date (2019-10-29) whichever comes first.

### Covariates

We included 5 major categories of covariates: demographics, laboratory tests related to CKD, diagnoses from medical history before index date, medications that are known to affect CKD related outcome, insurance status and smoking status. Demographics includes age, gender, race were collected at baseline (index date). Race categories included White, Black, Asian, and other races. We used White race as the reference group. Insurance status categories included Medicaid, self-pay, Medicare, commercial insurance. Commercial insurance was used as the reference group. Laboratory test included hemoglobin A1C (HbA1c), high density lipoprotein cholesterol (HDL), total cholesterol (TOTCHL), serum creatinine, body mass index (BMI). Medications include angiotensin-converting enzyme (ACE) inhibitors, aldosterone receptor antagonists (ARA), angiotensin II receptor blocker (ARB), antiplatelet drugs, beta-blockers, calcium channel blockers, diuretics, statins, and other lipid modifying drugs (loop and thiazide diuretics). Medical history includes cardiovascular disease (CVD), congestive heart failure (CHF), hypertension, vascular disease, vascular complication of diabetes (including skin ulcer), diabetic neuropathy, diabetic oculopathy, dyslipidemia, and other diabetic complications (diabetic nephropathy, nephrotic syndrome, nephritis/nephropathy, lower extremity amputations). For CKD hospitalization outcome, prior CKD hospitalization is also included as a covariate.

### Data cleaning

We included the medical history and medication use covariates for any time before the index date. For lab tests, we used the measurement closest to the index date in the prior 2 years. We treated All the lab tests (HbA1c, HDL, TOTCHL, BMI, serum creatinine) as continuous variables. We excluded extreme values that were likely to be erroneous values (e.g. HDL > 100 mg/dL, BMI< 8 kg/m, BMI> 100 kg/m, HbA1c>20% (195 mmol/mol), TOTCHL > 1000 mg/dL) from the study. A patient may not have any laboratory test during our 2-year searching time window. In addition, some patients also have missing values in race and insurance status in EHR. To deal with the missing data, we used multiple imputation by chained equation ^13^. In our analysis, we used predictive mean matching as the imputation method for continuous variables and polytomous regression imputation for unordered categorical data. We created 10 multiple imputed datasets. For each imputation, we set the number of iterations as 20. We fit cox regression model on each imputed dataset. We then used Rubin’s rule to pool coefficient estimates from 10 cox regression models ^14^.

### Statistical analysis

We generated a simple statistical summary and stratified patients by second-line ADM medication. For categorical variables, we conducted Chi-square test for differences among second-line ADM classes. For continuous variables, we performed t-test to test the differences among second-line ADM classes at baseline. To evaluate the association of ADM drug classes and three different CKD-related outcomes, we developed a series of cox proportional hazard regression models. In our baseline model, we included only ADM class variables. In our basic demographic model, we included both ADM class and basic demographic information. In our demographic-medical history model, we included ADM class variables, basic demographics, and medical history. In the full model, we included all the mentioned variables in the Covariates section. Cox regressions were conducted using ‘survival’ package in R, version 3.6.0. Multiple data imputation was performed using ‘MICE’ package in R, version 3.6.0. Estimate pooling from 10 imputed datasets were performed using ‘Hmisc’ in R, version 3.6.0. Descriptive data statistics were generated using ‘TableOne’ module in python, version 3.7.3.

### Sensitivity analysis

We conducted a sensitivity analysis to evaluate the robustness of our association findings. In the sensitivity analysis, we excluded patients with missing race, insurance status, HbA1c, total cholesterol, BMI, SBP, eGFR, and serum creatinine instead of imputing the missing data. The same cox regression models were performed to assess the association of ADMs with the three above mentioned outcomes.

## Results

We identified 3403 patients in our final cohort who started with Biguanides (mainly metformin) as their first ADM and used either a DPP-4 inhibitor, SGLT-2 inhibitor, GLP-1RA, TZD, SU, or an insulin as their second line medication. Table 1 shows the baseline characteristics of patients stratified by second-line ADM classes. For CKD incident outcome, we further excluded patients who had CKD diagnosis before the index date, which left us 3216 patients in the final cohort for CKD incidence outcome (see Figure 1).

Among the 3403 patients, overall, the mean age of the population is 59.8 (S.D = 12.0). Among these patients, 125 (3.67%) patients have missing data in race. For these who have race information, 2408 (73.5%) were white, 458 (14.0%) black, 178 (5.4%) Asian, 234 (7.1) % classified as other races; 1853 (54.5%) patients were male, and 1550 (45.5%) were female. There are 44 (1.29%) patients with missing insurance. Among the patients with insurance, 113 (3.5%) were self-pay, 1706 (50.8%) had commercial insurance, 128 (3.8%) were Medicaid, 1407 (41.9%) were Medicare. For laboratory test, there were 391 (11.49%) patients with missing HDL-C, 129 (3.79%) patients with missing HbA1c, 18 (0.5%) patients with missing BMI, and 462 (13.6%) patients with missing TOTCHL. For those patients with available laboratory values, the mean of HDL-C is 46.0 mg/dL (S.D. = 12.3), the mean of HbA1c is 7.4% (57 mmol/mol) (S.D. = 1.5), the mean of TOTCHL is 166.5 mg/dL (S.D. = 40.4), the median of serum creatinine is 79.58 umol/L ([Q1,Q3] = [61.89-88.42]). Among the 3403 patients, 1192 (35.0%) patients had DPP-4 inhibitors as second line ADM, 208 (6.1%) patients had GLP-1RA as second line ADM, 215 (6.3%) patients had insulin as second ADM, 1348 (39.6%) used SU as second line ADM, 355 (10.4%) patients had SGLT-2 inhibitor as second line ADM and 85 (2.5%) patients had TZD as second line ADM.

During the 5-year follow up, 232 of 3216 patients (7.2%) had incident CKD with median length of follow-up of 3.23 years. Out of 3403 patients, 109 (3.2%) patients had CKD hospitalization with median length of follow-up of 3.31 years. Figure 2 and Figure 3 showed the unadjusted Kaplan Meier curves for incident CKD and CKD hospitalization outcomes, respectively.

**Figure 2.**
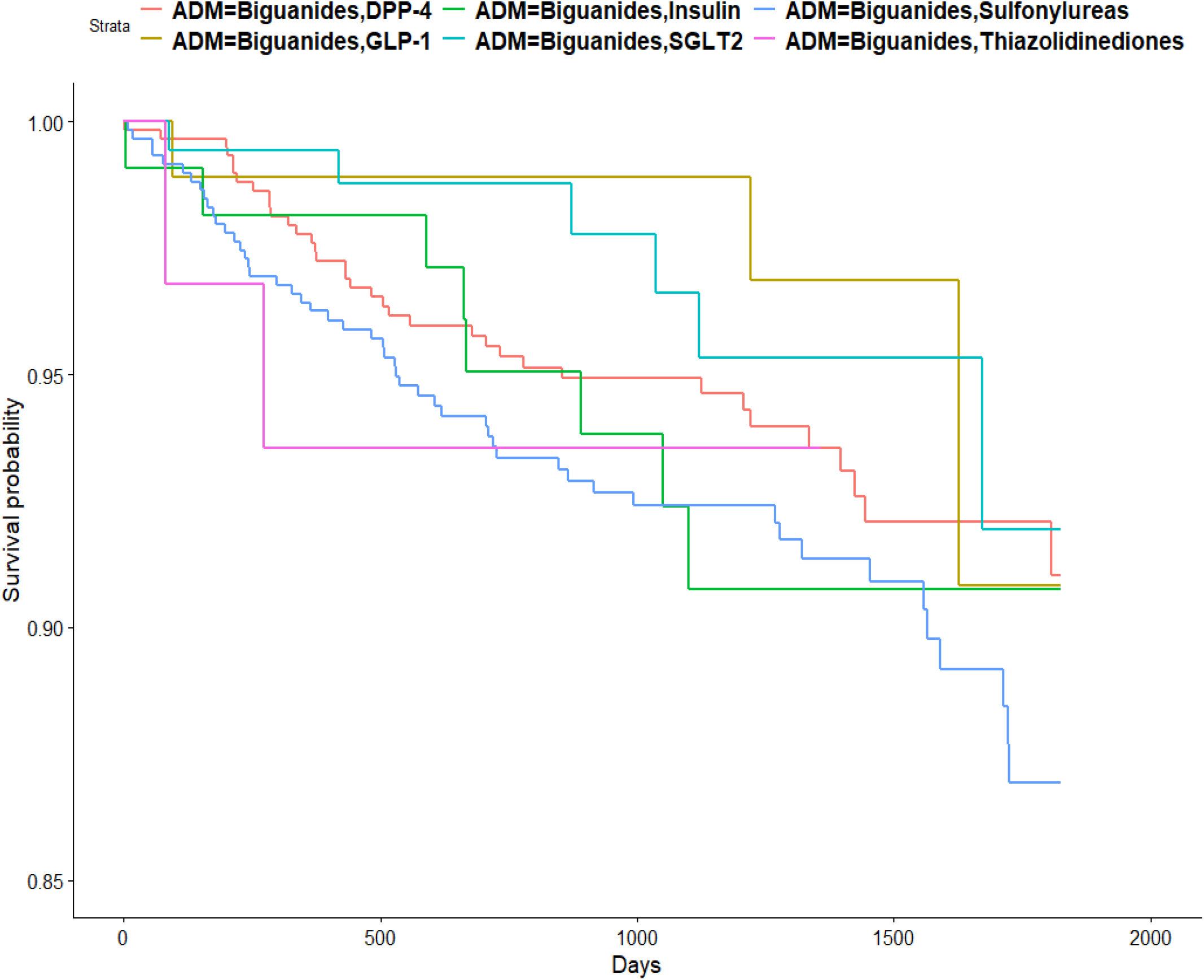
Unadjusted Kaplan Meier curve for incident CKD in different ADM groups. Abbreviations: CKD, chronic kidney disease; DPP4, dipeptidyl peptidase 4 inhibitors; GLP1RA, glucagon-like peptide receptor agonists; SGLT2 inhibitor, sodium-glucose cotransporter 2 inhibitor; TZD, Thiazolidinediones.

**Figure 3.**
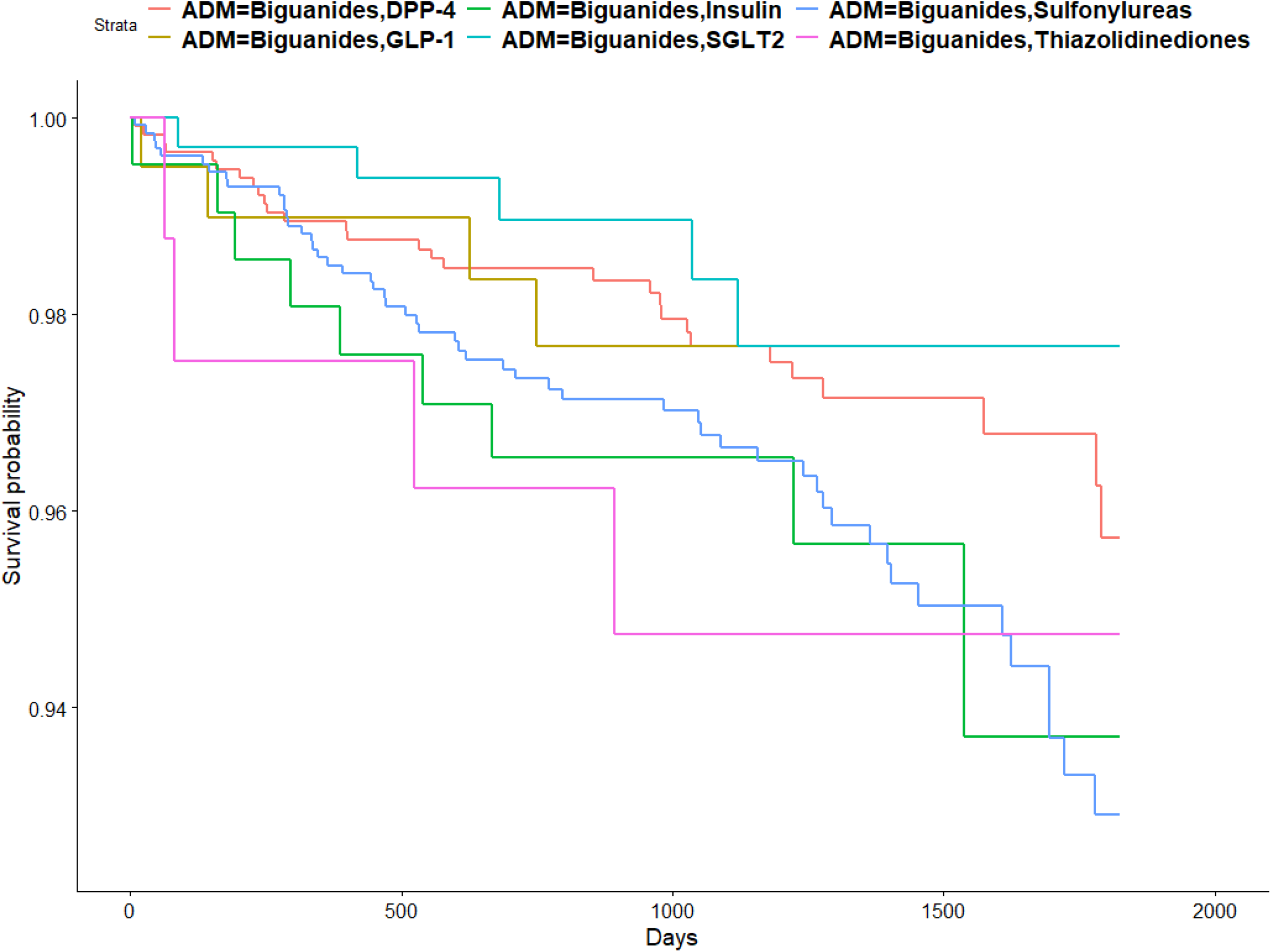
Unadjusted Kaplan Meier curve for CKD hospitalization in different ADM groups. Abbreviations: CKD, chronic kidney disease; DPP4, dipeptidyl peptidase 4 inhibitors; GLP1RA, glucagon-like peptide receptor agonists; SGLT2 inhibitor, sodium-glucose cotransporter 2 inhibitor; TZD, Thiazolidinediones.

### Risk for incident CKD and CKD hospitalization outcome in primary analysis

Table 2 showed the number of events and hazard ratio of incident CKD outcome for each second-line ADM class using SU as reference group in a fully adjusted model in the primary analysis (see Supplemental Table 3, 4 for hazard ratio in baseline model, basic demographics model, basic demographics/medical history model in primary analysis)^7^. For CKD incidence, both SGLT-2 inhibitor (HR, 0.43; 95% CI, [0.22;0.87]; P=0.02) and DPP-4 inhibitor (HR, 0.71; 95% CI, [0.53;0.96]; P=0.03) are significantly associated with lower CKD incidence event. GLP-1RA was associated with reduced risk for CKD incidence (HR, 0.52; 95% CI, [0.21;1.29]; P=0.16), but the P-value is not significant. For CKD hospitalization outcome, DPP-4 is significantly associated with lower risk for CKD hospitalization (HR, 0.60; 95% CI, [0.37;0.96]; P=0.03). Use of the other ADMs also did not show significant difference in CKD hospitalization outcome compared to use of SU.

**Table 2.**
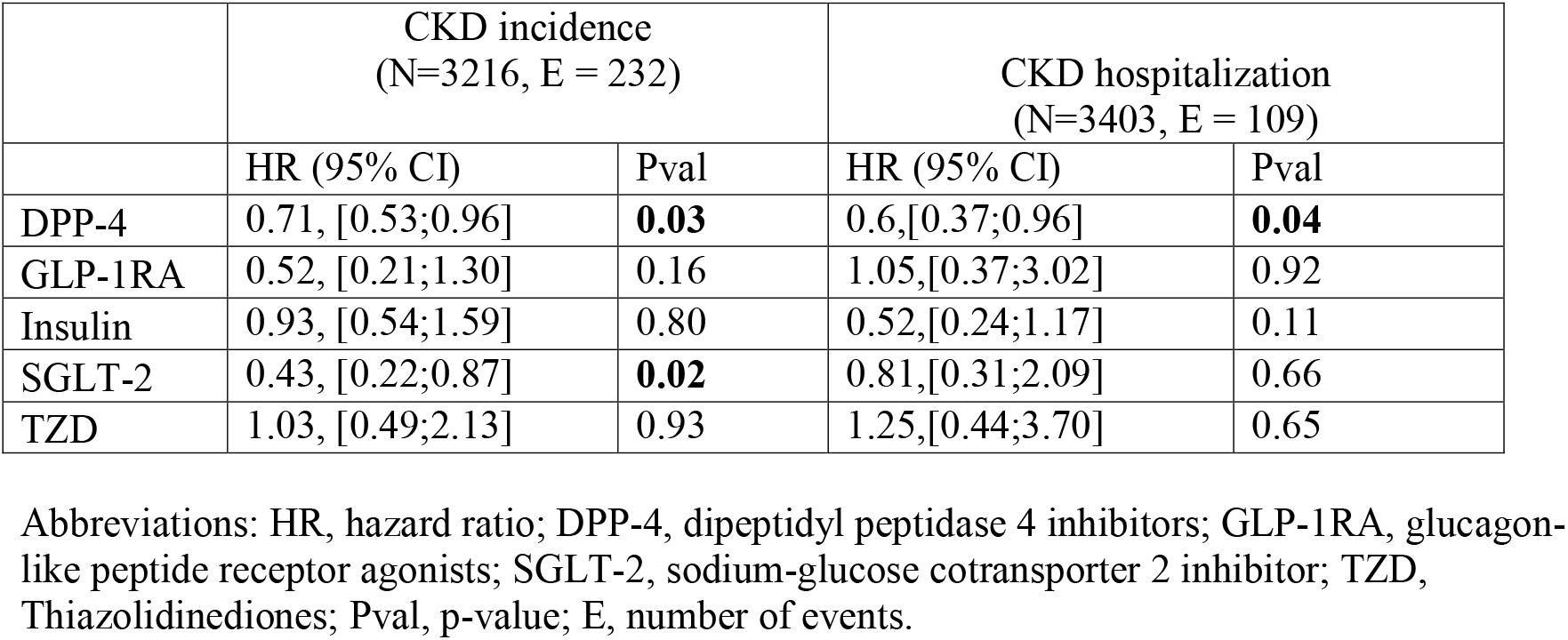
Hazard ratio in the fully adjusted cox regression model in primary analysis.

**Table 3.** Hazard ratio in the fully adjusted cox regression model in sensitivity analysis.

|  | CKD incidence<br>(N=2364, E=159) |  | CKD hospitalization<br>(N=2499, E =75) |  |
| --- | --- | --- | --- | --- |
|  | HR (95% CI) | Pval | HR (95% CI) | Pval |
| DPP-4 | 0.75; [0.52;1.08] | 0.12 | 0.75;[0.42;1.32] | 0.32 |
| GLP-1 | 0.53; [0.19;1.48] | 0.23 | 0.94;[0.26;3.43] | 0.92 |
| Insulin | 0.72; [0.37;1.43] | 0.35 | 1.00; [0.39;2.57] | 1.00 |
| SGLT-2 | 0.42; [0.19;0.92] | **0.03** | 1.35;[0.50;3.66] | 0.56 |
| TZD | 1.15; [0.49;2.71] | 0.75 | 0.9; [0.21;3.90] | 0.89 |

Abbreviations: HR, hazard ratio; DPP-4, dipeptidyl peptidase 4 inhibitors; GLP-1RA, glucagon-like peptide receptor agonists; SGLT-2, sodium-glucose cotransporter 2 inhibitor; TZD, Thiazolidinediones; Pval, p-value; E, number of events.

### Sensitivity analysis

Table 3 showed hazard ratios for ADMs relative to use of SU in the sensitivity analysis in fully adjusted model (see Supplemental Table 5, 6 for hazard ratio in baseline model, basic demographics model, basic demographics/medical history model in sensitivity analysis)^7^. Our sensitivity analyses included only those patients with complete data. For the incident CKD event, after excluding patients with missing race, insurance status, HbA1c, insurance status, HDL, total cholesterol, BMI, baseline serum creatinine, and baseline eGFR, 2364 patients remained. Our analysis showed that in the fully adjusted model, SGLT-2 inhibitor use (HR, 0.42; 95% CI, [0.19, 0.92], P=0.03) was significantly associated with lower incidence of CKD. DPP-4 inhibitor use (HR, 0.75; 95% CI, [0.52;1.08]; P=0.12) was associated with lower CKD incidence but the P value was not statistically significant. For the CKD hospitalization outcome, after excluding patients with missing values, there were 2499 patients left. we did not observe any significant association between a particular second line ADM class relative to SU.

## Discussion

Randomized clinical trials and observational studies are two major ways to assess associations between medication and corresponding clinical outcomes ^15^. Although randomized clinical trials are rigorously designed, they have several limitations in testing the association between ADMs and renal outcomes among type 2 diabetes patients. First, the major randomized clinical trials compared ADM’s effect to a placebo group. To date the ongoing Glycemia Reduction

### Approaches in Diabetes

A Comparative Effectiveness Study (GRADE) (54) is the only clinical trial (finished or ongoing) that we are aware of that evaluates the comparative effectiveness of newer ADMs vs older ADMs on glucose control ^16^. Further, most clinical trials treated second line ADMs as a monotherapy ^3,17^. Therefore, it is unclear how the effect changes when the ADMs are used as add-on therapy on top of metformin, which is the most common case in the primary care setting. Secondly, patients in clinical trials are better positioned to adhere to medication and therapy than in real clinical settings. Therefore, the results drawn from clinical trials may not apply to real clinical settings. Observational studies using EHR data like ours could potentially close these gaps by providing evidence from the real-world data of general populations.

In this study, we used EHR data to investigate the association of second-line ADMs with renal outcomes among 3403 type 2 diabetes patients who initiated treatment with metformin. We found that both the use of SGLT-2 inhibitor and DPP-4 inhibitor showed significant association with less incident CKD as compared to use of SU as a second-line medication. However, in the sensitivity analysis, only SGLT-2 inhibitor remained significantly associated. For the CKD hospitalization event, DPP-4 inhibitor appeared protective for CKD hospitalization in the primary analysis, but the association was not statistically significant in the sensitivity analysis. The above observations are largely in line with results from placebo-controlled cardiovascular outcomes trials, which showed empagliflozin, canagliflozin, dapagliflozin, and Linagliptin all had beneficial effects on indices of CKD ^18,19^. For example, the CREDENCE (Canagliflozin and Renal Events in Diabetes with Established Nephropathy Clinical Evaluation) trial showed that type 2 diabetes patients with prior albuminuria assigned canagliflozin (a SGLT-2 inhibitor) had lower risk of composite renal events compared to placebo group ^18^. Cooper et al. found Linagliptin (a DPP-4 inhibitor) significantly reduced the risk of kidney disease events compared to placebo among type 2 diabetes patients ^19^. Groop et al ^20^ found linagliptin on top of renin-angiotensin-aldosterone system significantly reduced albuminuria among patients with type 2 diabetes and renal disfunction. We did not find statistically significant difference between GLP-1RA and SU, Insulin and SU, TZD and SU in either primary analysis or sensitivity analysis regarding CKD incidence and CKD hospitalization event. For GLP-1RA, previous studies showed inconclusive evidence of its effect in reduction of renal disease ^21^. The LEADER ^21^ study showed GLP-1RA had benefit in reduction of new onset albuminuria while the LIRA-RENAL study showed no improvement in urine ACR, and eGFR change among renal impaired T2DM patients compared to the placebo group ^22^. For TZD vs SU, Insulin vs SU, we did not find evidence of one is superior to the other in renal disease outcome in the literature. Our study has several strengths. First, our clearly defined exposure time starting from first prescription of metformin, with additional prescription of a second line ADM within a year: this ensured comparison of patients who were in a clinically similar diabetes stage. Second, we only included patients who started ADMs after 2013-03-01 which is the time when the first SGLT-2 inhibitor was approved by FDA for use. This ensures that patients using different second-line ADMs have comparable follow-up times and avoids the immortal time bias ^23^. Third, we used a rigorously defined patient cohort: we excluded any patients who had type 1 diabetes diagnosis or gestational diabetes. In addition, we only included patients who had 3 continuous years of ADM prescriptions since their first ADM exposure. This makes sure that each patient has sufficient and comparable data depth. Last while there could be confounders that were not included in the study, our list of covariates is relatively comprehensive based on a review of the literature. This list contains basic demographic information, laboratory tests, and a wide range of diagnoses that might not only be associated with renal disease, but also suggest disease severity.

Our study has several limitations. Firstly, for simplicity, we could not distinguish between a switch of medication from an add-on medication. For example, patients who have a DPP-4 inhibitor as the second medication in their sequence could either: 1) have switched entirely from metformin to a DPP-4 inhibitor, or 2) began using metformin and a DPP-4 inhibitor together. Secondly, while we controlled for as many variables/confounders as we could, it is possible that there remain other covariates we did not capture, which could affect prescription bias or are still associated with the renal outcomes. Therefore, conclusions drawn from our study should be interpreted with caution. Thirdly, as EHR data is primarily designed for administrative purpose, it is well known that EHR’s have missing data issues. Because this is a single site study, patients may have sought care elsewhere. Therefore, it is likely that there were data points that we were unable to capture in this single site study. Additionally, this was a single center study of an academic hospital in a major US city, limiting its generalizability to other populations. Finally, diabetes duration is an important factor that affects renal outcomes. In our study, we did not control diabetes duration as a covariate because it was unavailable in our dataset. That said, we collected medication sequences at first exposure to ADMs, suggesting a clinically recent disease onset time. This makes the impact of not being able to include diabetic duration minimum.

## Conclusion

In conclusion, our study assessed the association of common second-line medications with renal outcomes as compared to SU’s from March 2013 through October 2019 using EHRs. Similar to previous studies, our results showed that SGLT-2 inhibitors were consistently associated with lower CKD incidence in both primary and sensitivity analyses and DPP-4 inhibitor was significantly associated with lower CKD incidence in primary analysis. We did not observe any statistical significance between Insulin and SU, TZD and SU in either primary analysis or sensitivity analysis regarding CKD incidence and CKD hospitalization event in our dataset. Unlike reported in Xie et al.’s study ^9^, We also did not observe significant benefit of GLP-1RA compared to SU in CKD hospitalization and CKD incidence, possibly due to small sample size. Additional research using multi-sites real-world data is needed to confirm the generalization of the results.

## Data Availability

The datasets generated and analyzed during the current study are not publicly available due to
protected patient information but are available from the corresponding author on reasonable request.

## Acknowledgements

The authors thank Raymond Kang (Northwestern University) for scientific discussions and technical assistance.

