## Supplemental Tables for "Use of real-world evidence data to evaluate the comparative effectiveness of second-line type 2 diabetes medications on chronic kidney disease"

Comparative Effectiveness of Second Line Anti-Diabetic Medication on Incidence of Chronic Kidney Disease

This appendix has been provided by the authors to give readers additional information about their work.

**Supplemental tables and figures**

**Table S1**. Diagnosis codes for type 2 diabetes

**Table S2.** Diagnosis codes for study variables

**Table S3.** Hazard ratio for CKD incidence outcome in primary analysis

**Table S4.** Hazard ratio for CKD hospitalization outcome in primary analysis

**Table S5.** Hazard ratio for CKD incidence outcome in sensitivity analysis

**Table S6.** Hazard ratio for CKD hospitalization outcome in sensitivity analysis

**Table S1.** Diagnosis codes for type 2 diabetes

| ICD 9 codes | 250.20, 250.30, 250.40, 250.50, 250.60, 250.70, 250.80, 250.90, 250.22, 250.32, 250.42, 250.52, 250.62, 250.72, 250.82, 250.92 |
| --- | --- |
| ICD 10 codes | E11.9, E13.01, E13.9, E11.00, E11.01, E13.10, E13.00 |
| SNOMED codes | 44054006, 197763012, 474213016, 200951011, 78158011, 48EB2F20-59A4-4676-A1C0-40880362224F, 359642000, 81531005, 719216001, 237599002, 199230006, 237627000, 9859006, 190331003, 703138006, 314903002, 190390000, 314902007, 190389009, 313436004 |

**Table S2.** Diagnosis codes for study variables

| **Covariates** | **ICD 9 codes** | **ICD 10 codes** |
| --- | --- | --- |
| CHF | 402.x1, 404.x1, 404.x3, 428.xx | I11.0, I13.0, I13.2, I50.x, I09.81 |
| Hypertension | 401.x-405.x | I10-13, I15-I16 |
| Hypoglycemic Events | 250.30, 250.32, 251.0, 251.1, 251.2 | E11.64x, E16.0, E16.1, E16.2 |
| Other diabetic complications | Diabetic Nephropathy: 250.40, 250.42  Nephrotic Syndrome: 581.x  Nephritis: 583.xx  Diabetic oculopathy: 250.50, 250.52, 362.0x, 366.41  Diabetic Cataracts: 366.41  Lower Extremity Amputations: V49.7x  Diabetic Retinopathy: 250.50, 250.52, 362.0x | Diabetic Nephropathy: E11.2x  Nephrotic Syndrome: N04.x  Nephritis: N05.x, N08  Diabetic oculopathy: E11.3x  Diabetic Cataracts: E11.36  Lower Extremity Amputations: Z89.4xx, Z89.5xx, Z89.6xx  Diabetic Retinopathy: E11.31 – E11.35 |
| Diabetic neuropathy | 250.60, 250.62, 357.2, 362.01-362.06 | E11.4x |
| Dyslipidemia | 272.0, 272.1, 272.2, 272.3, 272.4 | E78.0 – E78.5, E78.00 |
| Tobacco Use | 305.1, V15.82 | F17.2x, Z87.891 |
| CVD | Stroke: 430, 431, 432.x, 433.x1, 434.x1, 435.x  PAD: 440.x, 441.x, 443.2x, 444.x, 445.x  IHD: 410.x, 411.x, 414.12 | Stroke: I63.x, I60.x, I61.x, I62.x, I63.xxx, G45.0-G45.2, G45.8, G45.9  PAD: I70.x, I71.x, I74.x, I75.x, I77.7x  IHD: I20.x, I21.x, I22.x, I23.x, I24.x, I25.42 |
| Vascular Disease | 440.x, 441.x, 442.x, 443.2x, 443.9, 444.x, 445.x | I70.x, I71.x, I72.x, I73.9, I74.x, I75.x, I77.7x |
| VCD | 250.70, 250.72, 607.84, 707.1x, 707.8, 707.9 | E11.5x, E11.62x, L97.xxx |
| **Renal outcomes** |  |  |
| End stage renal disorder | 585.6 | N18.6 |
| Chronic kidney disease | 285.21, 403.x, 404.x, 582.x, 585.x, 586.x | D63.1, E11.22, I12.x, I13.x, N03.x, N18.x |

Abbreviations: CHF, congestive heart failure; CVD, cardiovascular disease; VCD: vascular complications of diabetes.

**Table S3.** Hazard ratio for CKD incidence outcome in primary analysis

| Sequence group | HR | HR 95% CI | P-value |
| --- | --- | --- | --- |
| Baseline model | | | |
| Biguanides, DPP-4 inhibitor | 0.64 | [0.48;0.86] | 0.00 |
| Biguanides, GLP-1RA | 0.28 | [0.11;0.67] | 0.00 |
| Biguanides, Insulin | 0.91 | [0.55;1.50] | 0.70 |
| Biguanides, SGLT-2 inhibitor | 0.31 | [0.16;0.60] | <0.001 |
| Biguanides, TZD | 1.00 | [0.49;2.04] | 0.99 |
| Basic demographics model | | |  |
| Biguanides, DPP-4 inhibitor | 0.73 | [0.54;0.98] | 0.03 |
| Biguanides, GLP-1RA | 0.45 | [0.18;1.10] | 0.08 |
| Biguanides, Insulin | 1 | [0.60;1.68] | 0.99 |
| Biguanides, SGLT-2 inhibitor | 0.43 | [0.22;0.86] | 0.02 |
| Biguanides, TZD | 1.06 | [0.52;2.17] | 0.87 |
| Basic demographics/medical history model | | | |
| Biguanides, DPP-4 inhibitor | 0.70 | [0.52;0.95] | 0.02 |
| Biguanides, GLP-1RA | 0.40 | [0.16;0.98] | 0.05 |
| Biguanides, Insulin | 0.81 | [0.48;1.37] | 0.44 |
| Biguanides, SGLT-2 inhibitor | 0.43 | [0.22;0.86] | 0.02 |
| Biguanides, TZD | 0.98 | [0.47;2.04] | 0.96 |
| Fully adjusted model | |  |  |
| Biguanides, DPP-4 inhibitor | 0.71 | [0.53;0.96] | 0.03 |
| Biguanides, GLP-1RA | 0.52 | [0.21;1.30] | 0.16 |
| Biguanides, Insulin | 0.93 | [0.55;1.59] | 0.80 |
| Biguanides, SGLT-2 inhibitor | 0.43 | [0.22;0.87] | 0.02 |
| Biguanides, TZD | 1.03 | [0.50;2.15] | 0.93 |

Abbreviations: HR, hazard ratio; CI, confidence interval; DPP-4 inhibitor, dipeptidyl peptidase 4 inhibitors; GLP-1RA, glucagon-like peptide receptor agonists; SGLT-2 inhibitor, sodium-glucose cotransporter 2 inhibitor; Thiazolidinediones, TZD

**Table S4.** Hazard ratio for CKD hospitalization outcome in primary analysis

| Variable | HR | HR 95% CI | Pval |
| --- | --- | --- | --- |
| Baseline model | | | |
| Biguanides, DPP-4 inhibitor | 0.57 | [0.36;0.89] | 0.01 |
| Biguanides, GLP-1RA | 0.49 | [0.18;1.34] | 0.16 |
| Biguanides, Insulin | 0.97 | [0.48;1.97] | 0.94 |
| Biguanides, SGLT-2 inhibitor | 0.38 | [0.15;0.95] | 0.04 |
| Biguanides, TZD | 1.02 | [0.37;2.82] | 0.96 |
| Basic demographics model | | | |
| Biguanides, DPP-4 inhibitor | 0.69 | [0.44;1.07] | 0.10 |
| Biguanides, GLP-1RA | 0.99 | [0.35;2.77] | 0.99 |
| Biguanides, Insulin | 1.05 | [0.52;2.15] | 0.88 |
| Biguanides, SGLT-2 inhibitor | 0.70 | [0.28;1.77] | 0.46 |
| Biguanides, TZD | 1.06 | [0.38;2.94] | 0.91 |
| Basic demographics/medical history model | | | |
| Biguanides, DPP-4 inhibitor | 0.56 | [0.35;0.91] | 0.02 |
| Biguanides, GLP-1RA | 0.96 | [0.34;2.71] | 0.93 |
| Biguanides, Insulin | 0.52 | [0.24;1.14] | 0.10 |
| Biguanides, SGLT-2 inhibitor | 0.85 | [0.33;2.16] | 0.73 |
| Biguanides, TZD | 1.27 | [0.45;3.61] | 0.66 |
| Fully adjusted model | | | |
| Biguanides, DPP-4 inhibitor | 0.60 | [0.37;0.96] | 0.03 |
| Biguanides, GLP-1RA | 1.05 | [0.37;3.02] | 0.92 |
| Biguanides, Insulin | 0.52 | [0.24;1.17] | 0.11 |
| Biguanides, SGLT-2 inhibitor | 0.81 | [0.31;2.09] | 0.66 |
| Biguanides, TZD | 1.25 | [0.44;3.70] | 0.65 |

Abbreviations: HR, hazard ratio; CI, confidence interval; DPP-4 inhibitor, dipeptidyl peptidase 4 inhibitors; GLP-1RA, glucagon-like peptide receptor agonists; SGLT-2 inhibitor, sodium-glucose cotransporter 2 inhibitor; Thiazolidinediones, TZD

**Table S5.** Hazard ratio for CKD incidence outcome in sensitivity analysis

| Variable | HR | HR 95% CI | P-value |
| --- | --- | --- | --- |
| Baseline model | | | |
| Biguanides, DPP-4 inhibitor | 0.68 | [0.48;0.97] | 0.03 |
| Biguanides, GLP-1RA | 0.35 | [0.13;0.97] | 0.04 |
| Biguanides, Insulin | 0.95 | [0.51;1.78] | 0.87 |
| Biguanides, SGLT-2 inhibitor | 0.35 | [0.16;0.75] | 0.01 |
| Biguanides, TZD | 1.19 | [0.52;2.73] | 0.68 |
| Basic demographics | | | |
| Biguanides, DPP-4 inhibitor | 0.76 | [0.54;1.09] | 0.14 |
| Biguanides, GLP-1RA | 0.53 | [0.19;1.47] | 0.22 |
| Biguanides, Insulin | 1.07 | [0.57;2.04] | 0.83 |
| Biguanides, SGLT-2 inhibitor | 0.46 | [0.21;1.00] | 0.05 |
| Biguanides, TZD | 1.13 | [0.49;2.61] | 0.77 |
| Basic demographics/medical history model | | | |
| Biguanides, DPP-4 inhibitor | 0.73 | [0.51;1.04] | 0.09 |
| Biguanides, GLP-1RA | 0.47 | [0.17;1.29] | 0.14 |
| Biguanides, Insulin | 0.85 | [0.44;1.64] | 0.62 |
| Biguanides, SGLT-2 inhibitor | 0.46 | [0.21;1.00] | 0.05 |
| Biguanides, TZD | 1.05 | [0.45;2.44] | 0.92 |
| Fully adjusted model | | | |
| Biguanides, DPP-4 inhibitor | 0.75 | [0.52;1.08] | 0.12 |
| Biguanides, GLP-1RA | 0.53 | [0.19;1.48] | 0.23 |
| Biguanides, Insulin | 0.72 | [0.37;1.43] | 0.35 |
| Biguanides, SGLT-2 inhibitor | 0.42 | [0.19;0.92] | 0.03 |
| Biguanides, TZD | 1.15 | [0.49;2.71] | 0.75 |

Abbreviations: HR, hazard ratio; CI, confidence interval; DPP-4 inhibitor, dipeptidyl peptidase 4 inhibitors; GLP-1RA, glucagon-like peptide receptor agonists; SGLT-2 inhibitor, sodium-glucose cotransporter 2 inhibitor; Thiazolidinediones, TZD

**Table S6.** Hazard ratio for CKD hospitalization outcome in sensitivity analysis

| Variable | HR | HR 95% CI | P-value |
| --- | --- | --- | --- |
| Baseline model | | | |
| Biguanides, DPP-4 inhibitor | 0.69 | [0.41;1.16] | 0.16 |
| Biguanides, GLP-1RA | 0.63 | [0.20;2.06] | 0.45 |
| Biguanides, Insulin | 1.16 | [0.49;2.74] | 0.74 |
| Biguanides, SGLT-2 inhibitor | 0.59 | [0.23;1.51] | 0.28 |
| Biguanides, TZD | 0.83 | [0.20;3.45] | 0.80 |
| Basic demographics | | | |
| Biguanides, DPP-4 inhibitor | 0.85 | [0.50;1.44] | 0.55 |
| Biguanides, GLP-1RA | 1.17 | [0.35;3.90] | 0.80 |
| Biguanides, Insulin | 1.21 | [0.50;2.93] | 0.67 |
| Biguanides, SGLT-2 inhibitor | 1.10 | [0.43;2.85] | 0.84 |
| Biguanides, TZD | 0.79 | [0.19;3.28] | 0.74 |
| Basic demographics/medical history model | | | |
| Biguanides, DPP-4 inhibitor | 0.65 | [0.37;1.15] | 0.14 |
| Biguanides, GLP-1RA | 1.03 | [0.30;3.57] | 0.96 |
| Biguanides, Insulin | 0.84 | [0.33;2.14] | 0.71 |
| Biguanides, SGLT-2 inhibitor | 1.30 | [0.49;3.48] | 0.60 |
| Biguanides, TZD | 0.92 | [0.21;3.98] | 0.92 |
| Fully adjusted model | | | |
| Biguanides, DPP-4 inhibitor | 0.75 | [0.42;1.32] | 0.32 |
| Biguanides, GLP-1RA | 0.94 | [0.26;3.43] | 0.92 |
| Biguanides, Insulin | 1.00 | [0.39;2.57] | 1.00 |
| Biguanides, SGLT-2 inhibitor | 1.35 | [0.50;3.66] | 0.56 |
| Biguanides, TZD | 0.90 | [0.21;3.90] | 0.89 |

Abbreviations: HR, hazard ratio; CI, confidence interval; DPP-4 inhibitor, dipeptidyl peptidase 4 inhibitors; GLP-1RA, glucagon-like peptide receptor agonists; SGLT-2 inhibitor, sodium-glucose cotransporter 2 inhibitor; Thiazolidinediones, TZD
